# Development of a highly sensitive point-of-care test to detect SARS-CoV-2 from saliva combining a simple RNA extraction method with colorimetric reverse transcription loop-mediated isothermal amplification detection

**DOI:** 10.1101/2020.11.08.20227702

**Authors:** Wataru Yamazaki, Yasufumi Matsumura, Uraiwan Thongchankaew-Seo, Yasuko Yamazaki, Miki Nagao

## Abstract

To diagnose COVID-19 patients in the field, a sensitive point-of-care test using saliva was developed. Using a heat block without centrifuge, the test took 45 minutes. Naked eye judgement with color change dye outperformed the reference standard, with a diagnostic sensitivity of 82.6% (19/23) and diagnostic specificity of 100% (21/21).

## Introduction

With progressing globalization, the worldwide spread of infectious diseases has accelerated. The new coronavirus infection (COVID-19) is a major public health concern (WHO, 2020). With the high burden and risk for infection among patients and healthcare workers, the advent of reverse transcription quantitative real-time polymerase chain reaction (RT-qPCR) testing as an alternative to nasopharyngeal swab sampling for diagnosing COVID-19 has increased the use of saliva (Azzi et al., 2020; Iwasaki et al., 2020; Nagura-Ikeda et al., 2020; To et al., 2020; Vogels et al., 2020; Williams et al., 2020; Wyllie et al., 2020).

Angiotensin-converting enzyme II (ACE2), the main receptor for SARS-CoV-2 entry into human cells, is highly expressed on the oral cavity mucosa, especially tongue epithelial cells (Iwasaki et al., 2020; Xu et al., 2020). In Japan, many COVID-19 cases have been found in metropolitan entertainment districts, where food, drink, and conversation are frequent. Saliva droplets containing SARS-COV-2 are a major vector for COVID-19 infection, making saliva a promising candidate for COVID-19 testing (Azzi et al., 2020; Nagura-Ikeda et al., 2020; Wyllie et al., 2020). While the viral load of SARS-CoV-2 in saliva samples is lower than in nasopharyngeal swab samples (Iwasaki et al., 2020; Williams et al., 2020), several researchers have reported comparable or better diagnostic sensitivity with saliva testing (Azzi et al., 2020; Nagura-Ikeda et al., 2020; To et al., 2020; Vogels et al., 2020; Wyllie et al., 2020).

The combined use of the direct-to-test addition by heating and RT-qPCR using a nasopharyngeal swab sample has proven successful due to reduced inhibitory effects (Esbin et. al, 2020). However, the saliva sample must undergo high-speed centrifugation at 20,000 *g* for 5-30 min following purification with a commercial RNA extraction kit to remove potential inhibitors (Azzi et al., 2020; Iwasaki et al., 2020; Nagura-Ikeda et al., 2020; To et al., 2020; Williams et al., 2020; Wyllie et al., 2020). This tedious RNA extraction process, which requires a well-equipped laboratory, is a major hurdle for performing sensitive point-of-care testing (POCT) for COVID-19 using saliva.

Reverse transcription loop-mediated isothermal amplification (RT-LAMP) is an attractive option for achieving more-sensitive POCT than an immunochromatographic assay because of its broader utility and portable real-time detector with an in-house buttery or combination use with a cost-effective heat block and colorimetric reagent (Augustine et al., 2020; Ecke et al., 2020; Hayashida et al., 2015; Park, et al., 2020).

Here, we developed a simple method for extracting RNA from saliva samples using semi-alkaline proteinase (SAP), a sputum homogenizer for tuberculosis examinations, and a subsequent simple heating step, with no need for centrifugation or RNA extraction. Further, we newly developed a triplex RT-LAMP approach using colorimetric readout in a heat block for the naked eye judgement. We preliminarily evaluated the POCT performance with 44 clinical saliva samples.

### The study

We used Primer Explorer V5 (primerexplorer.jp/lampv5e/index.html) to design three primer sets targeting the ORF 1ab, S, and ORF 7a regions to simultaneously detect SARS-CoV-2 with high sensitivity and robustness. To identify highly conserved nucleotide sequences with low similarity to SARS-CoV-1 sequences, we performed multiple sequence alignment using Clustal Omega (www.ebi.ac.uk/Tools/msa/clustalo) of 490 whole-genome SARS-Cov-2 sequences from the DDBJ/EMBL/GenBank databases (www.ncbi.nlm.nih.gov/nucleotide/) submitted by worldwide researchers from five continents. The details of each primer and in-house reagent are shown in S-Tables 1-3. Artificially synthesized SARS-CoV-2 sequences (Eurofins, Tokyo, Japan) corresponding to the three target regions and RNA extracted from healthy human saliva were used as positive and negative controls, respectively, for the triplex RT-LAMP format.

From May to July 2020, we collected at least 1 ml of saliva samples from 44 clinical patients suspected of having COVID-19 infection at Kyoto University Hospital or Kyoto City Hospital. The saliva samples were mixed with 3 strength-volume of SAP (Semi-Alkaline Proteinase, Suputazyme; Kyokuto Pharmaceutical Industrial, Tokyo, Japan) manually or using a vortex mixer for 15 sec. After incubation for 15 min at room temperature to dissolve the saliva components, each of the saliva-SAP mixtures (SS mixes) were added to a 1.5-ml microcentrifuge tube. For RNA extraction, 140 μl of each SS mix was eluted into 60 μl using a QIAamp Viral RNA Mini Kit (Qiagen, Hilden, Germany). RT-qPCR using the N2 primers and probe was performed with 5 μl of the extracted RNA in 20 μl of reaction mixture in LightCycler 480 System II (Roche, Basel, Switzerland) according to the protocol recommended by the National Institute of Infectious Diseases, Japan (Shirato et al., 2020). Ten-fold serial dilutions (5 to 5×10^6^ copies/reaction) of the synthesized RNA containing the target sequence (Shirato et al., 2020) were used to calculate the viral loads in clinical samples.

In parallel, for POCT, the remaining SS mix was heated at 95 °C for 5 min, and then the tube was centrifuged using a dry-cell battery-powered tabletop centrifuge for approximately 5 sec or swung manually to remove the aqueous droplet on the tube lid. Using 50 µl of RT-LAMP reaction mixtures (including 10 μl of template), the RT-LAMP reaction was monitored using a LightCycler 480 System II (Roche) at 63 °C for 25 min. Subsequent procedures were completed without interruption, or else the SS mix was stored at -80 °C until used. The limit of detection (LOD) of triplex RT-LAMP was defined as the lowest concentration at which 95% of positive samples were detected using a 2-fold series of phosphate-buffered saline dilutions in maximum 20 replicates of 50,000, 25,000, 12,500, and 10,000 copies/ml of heat-inactivated SARS-CoV-2 (ATCC VR-1986HK; American Type Culture Collection, Manassas, VA, USA) (Table 2). The dilutions were heated at 95 °C for 5 min. After removal of the aqueous droplet on the tube lid, 10 μl of the template was applied to 50 µl of RT-LAMP reaction mixture, as described above.

**Table 1.**
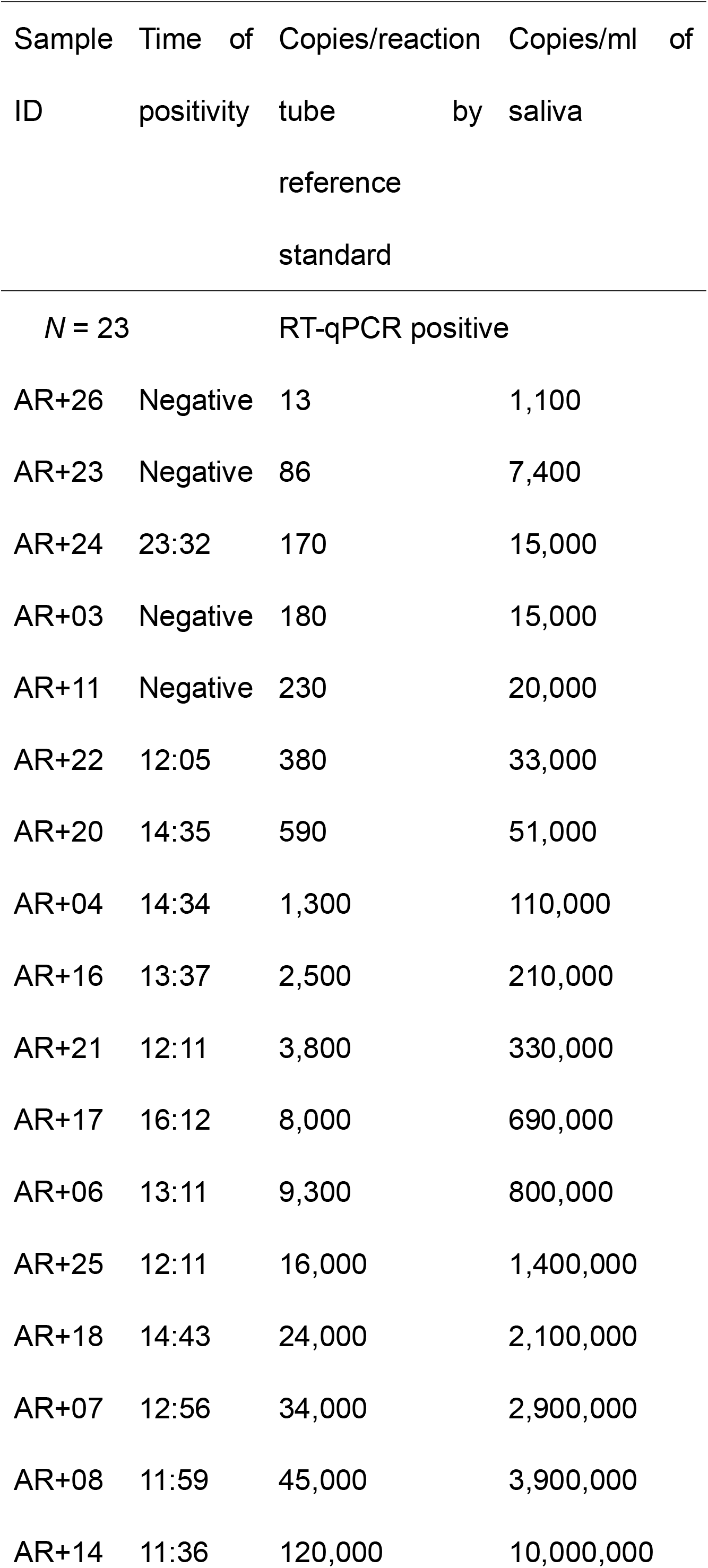
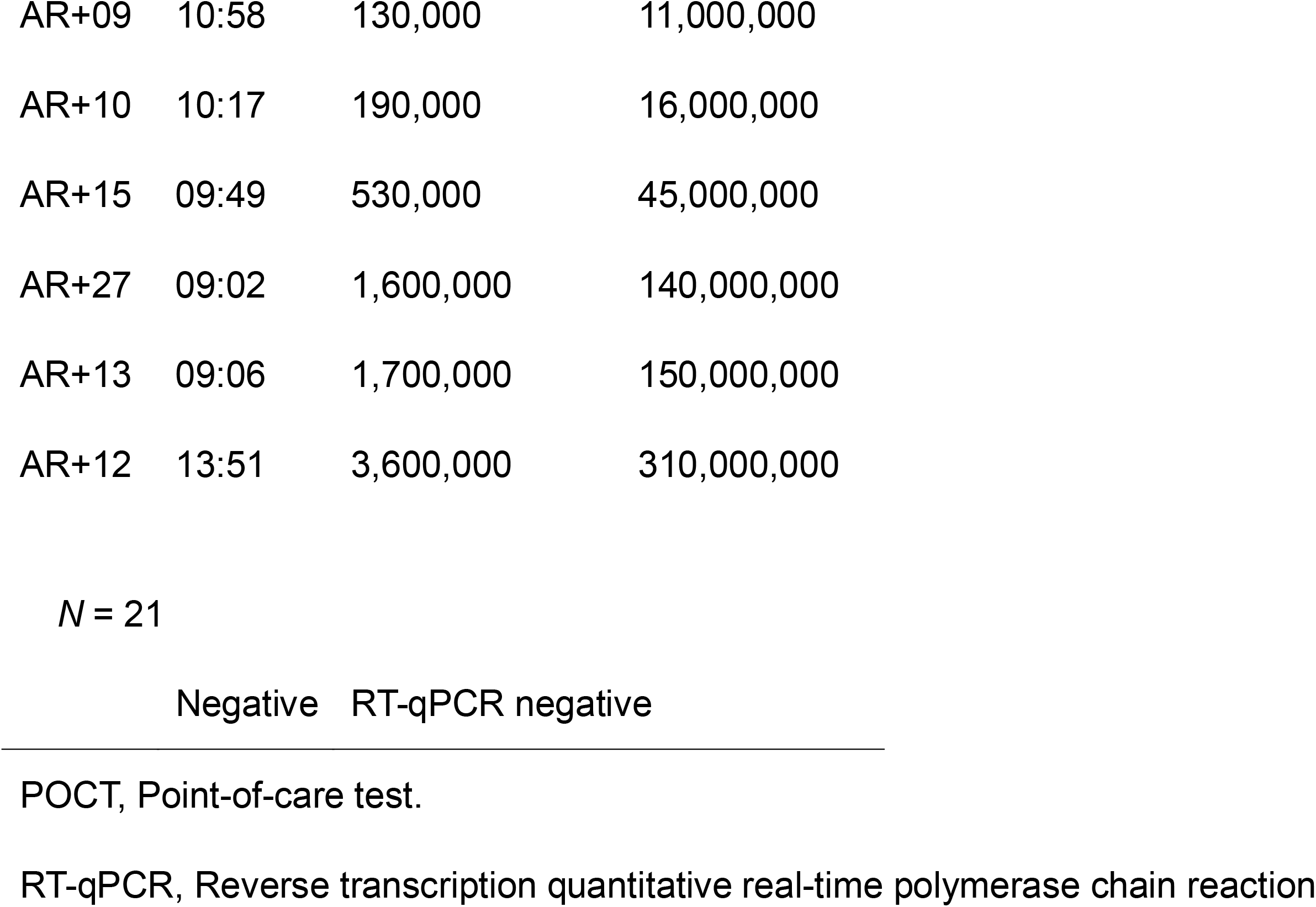
Results of POCT and reference standard in 44 clinical patients.

**Table 2.**
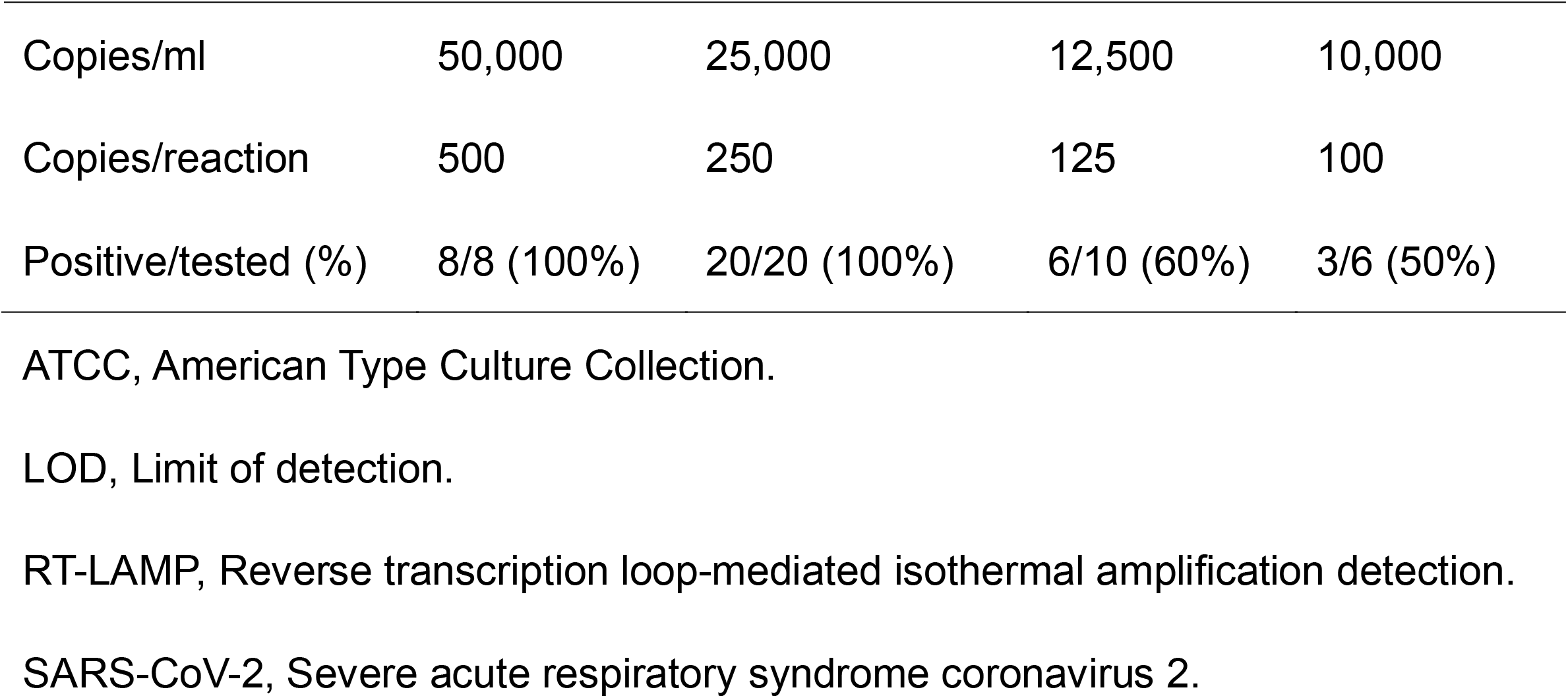
LOD determination of the triplex RT-LAMP with the heat-inactivated SARS-CoV-2, ATCC VR-1986HK in phosphate-buffered saline.

|  |  |  |  |  |
| --- | --- | --- | --- | --- |
| Copies/ml | 50,000 | 25,000 | 12,500 | 10,000 |
| Copies/reaction | 500 | 250 | 125 | 100 |
| Positive/tested (%) | 8/8 (100%) | 20/20 (100%) | 6/10 (60%) | 3/6 (50%) |

ATCC, American Type Culture Collection.
LOD, Limit of detection.
RT-LAMP, Reverse transcription loop-mediated isothermal amplification detection.
SARS-CoV-2, Severe acute respiratory syndrome coronavirus 2.

Compared to reference standard, the diagnostic sensitivity and specificity were 82.6% (19/23; 95% confidence interval [CI], 61.2%-95.1%) and 100% (21/21, 95% CI, 83.8%-100%) for the new POCT, which took 45 min from starting crude RNA extraction to endpoint readout with the naked eye (Table 1, Figure 1). As shown in Table 1, 18 clinical samples exceeding 380 copies/reaction (>33,000 copies/ml of saliva) by reference standard measurement were positive on POCT between 9:02 and 16:12 for amplification. Three samples showing low copy numbers at 170-230 copies/reaction (15,000-20,000 copies/ml of saliva) on RT-qPCR were only 1 positive at 23:32 and 2 false negatives in POCT, which were interpreted as borderline samples near the LOD of POCT. Another 2 samples showing lower copy numbers at 13-86 copies/reaction (1,100-7,400 copies/ml of saliva) were both false negatives, possibly due to below the detection power of the POCT. The result obtained by the LOD determination was the value of 250 copies/reaction (25,000 copies/ml) (Table 2).

**Figure 1.**
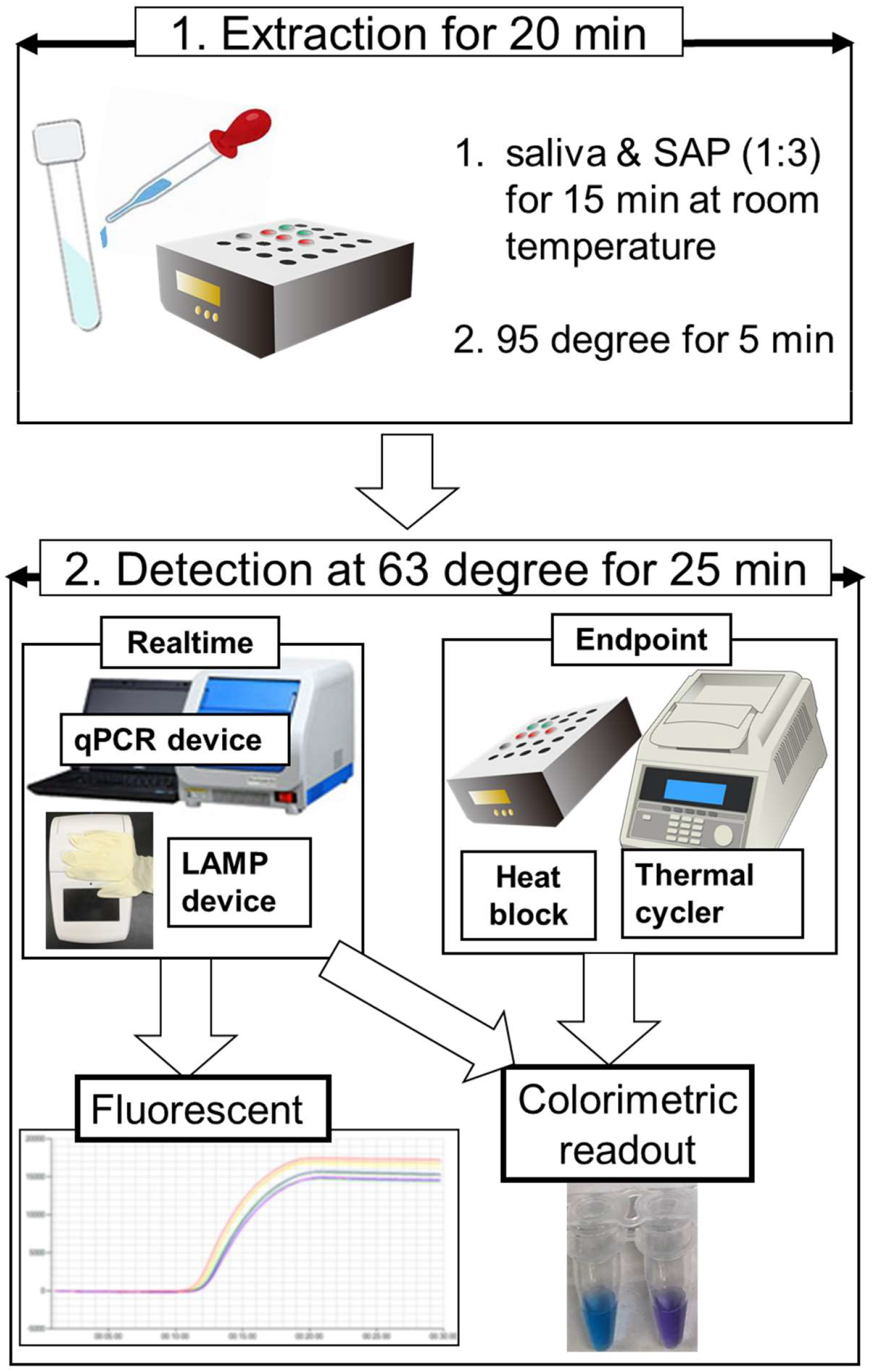
Concept of POCT using a combination of simple RNA extraction and LAMP devices.

## Conclusion

Our newly developed POCT approach achieved simple RNA extraction and constant RT-LAMP without the need for high-speed centrifugation, nucleic acid extraction kits, or qPCR equipment. This POCT does not require expensive equipment and performs comparably to an existing reference standard. This POCT can thus be used for drive-through testing and simple inspection stations in a field setting, helping reduce the risk of infection by simplifying and accelerating testing for COVID-19.

## Data Availability

The data that support the findings of this study are available from the corresponding author WY.

## Acknowledgements

This research was supported by AMED under Grant Number JP20he0622031.

## Ethics approval

The Ethics Committee of Kyoto University Graduate School and the Faculty of Medicine approved this study (R2379) and waived the need for obtaining informed consent from each patient.

## First author biographical sketch

Wataru Yamazaki is a veterinarian and microbiologist. He specializes in food microbiology and infectious diseases for both humans and animals and has been particularly involved in the development of LAMP and other molecular-based testing methods as well as POCT for fieldwork for over 15 years.

## Figure legend

**Technical Appendix. S-Table 1.**
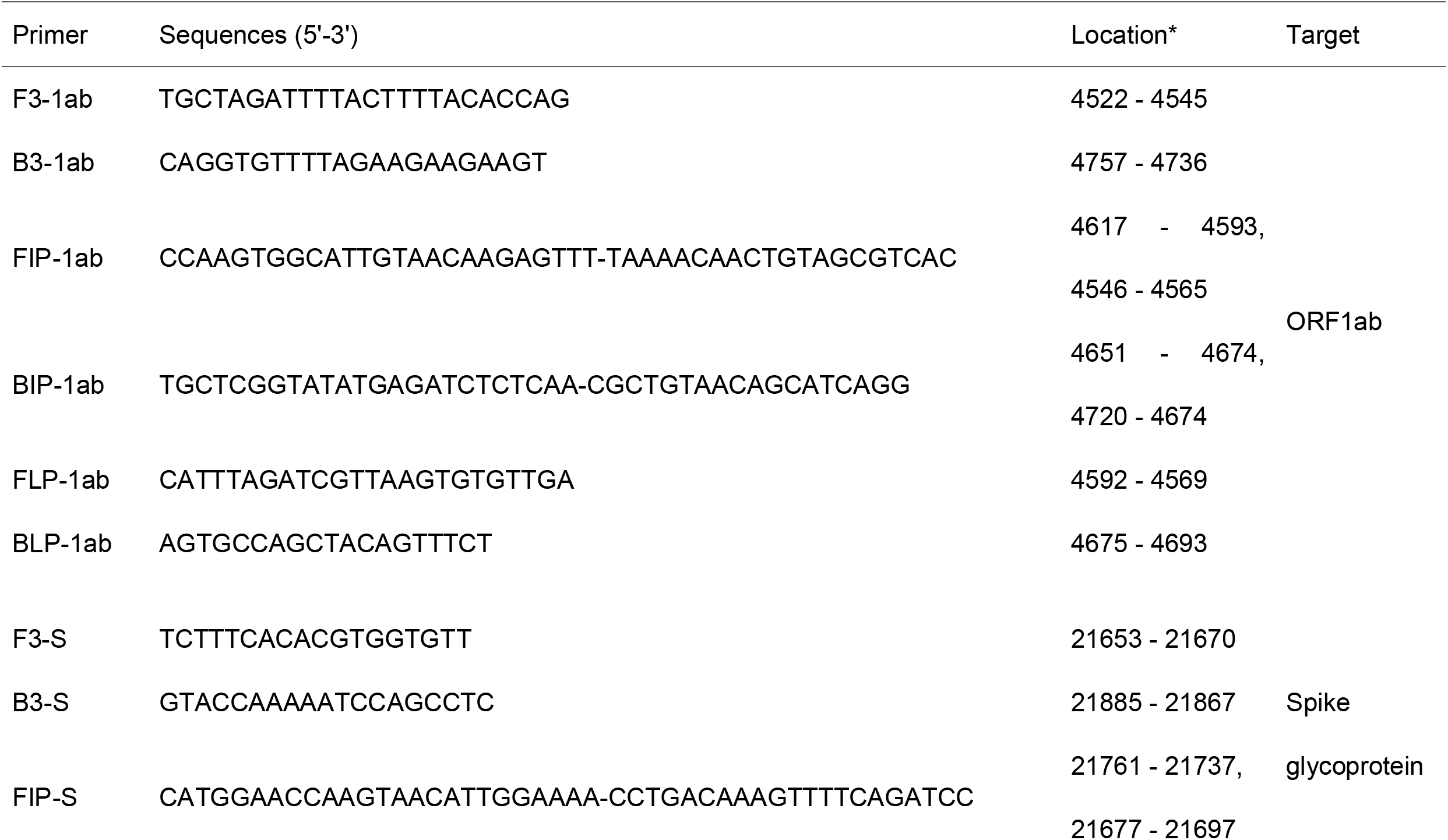
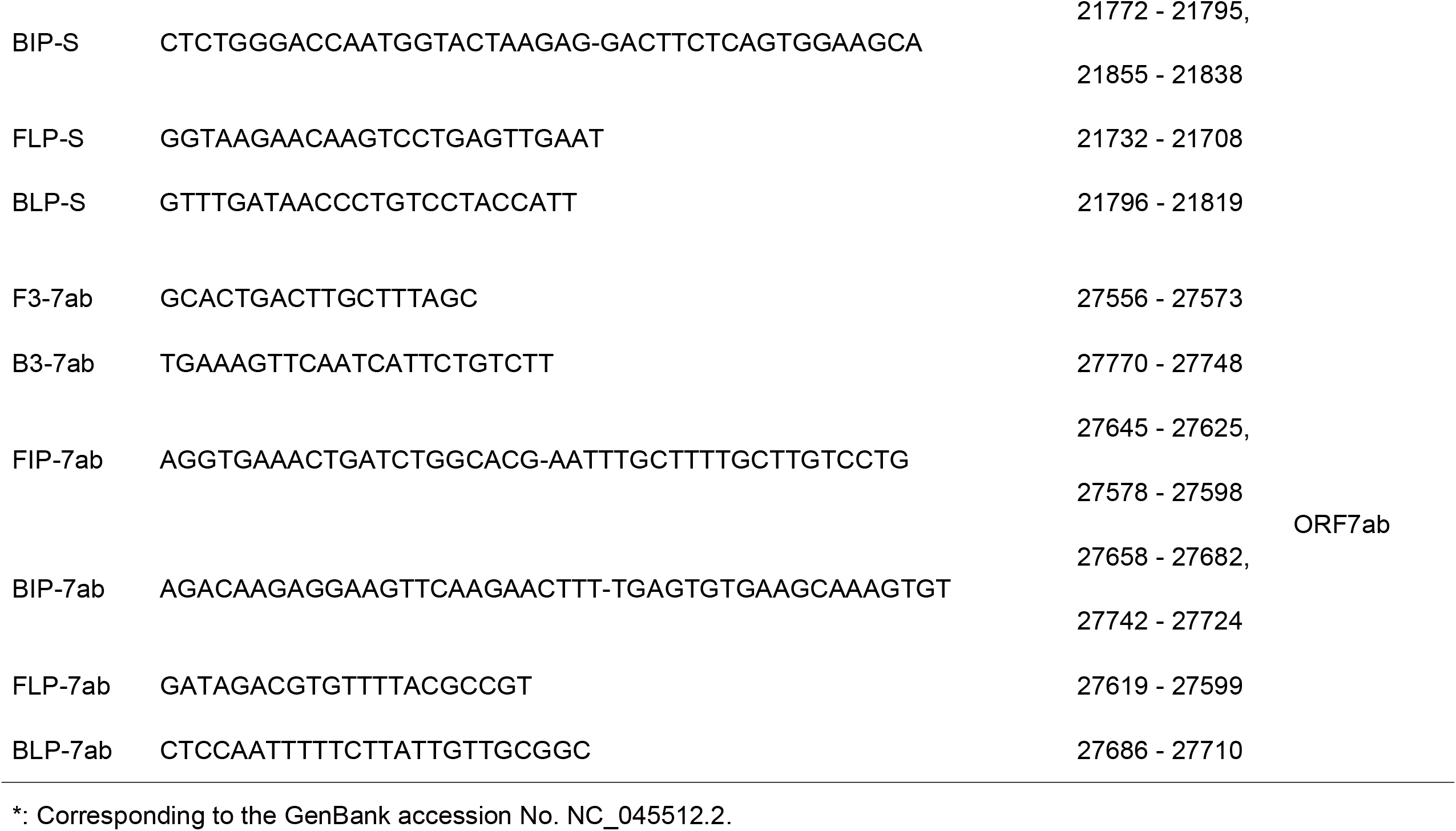
Primers used for RT-LAMP assay.

**S-Table 2.**
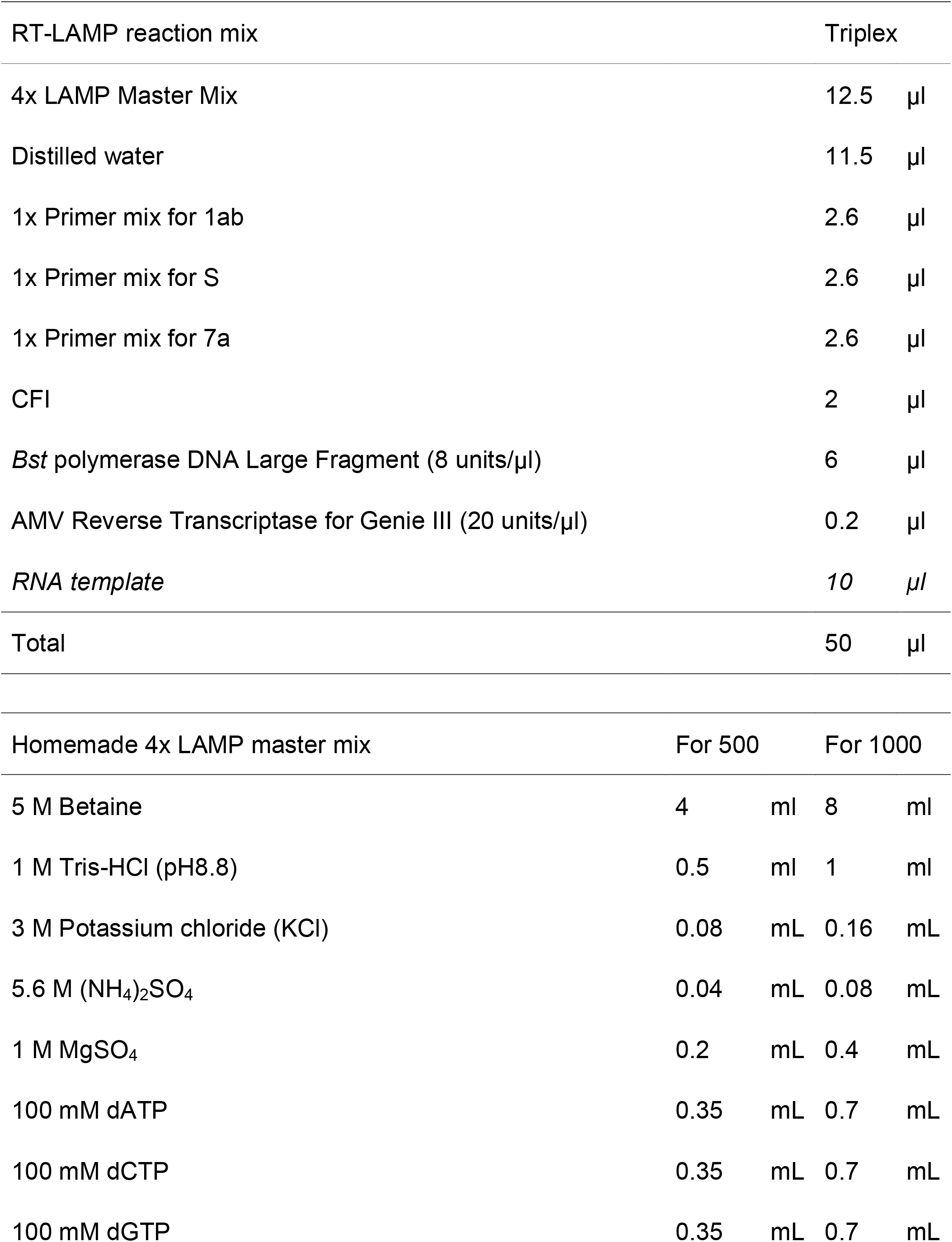
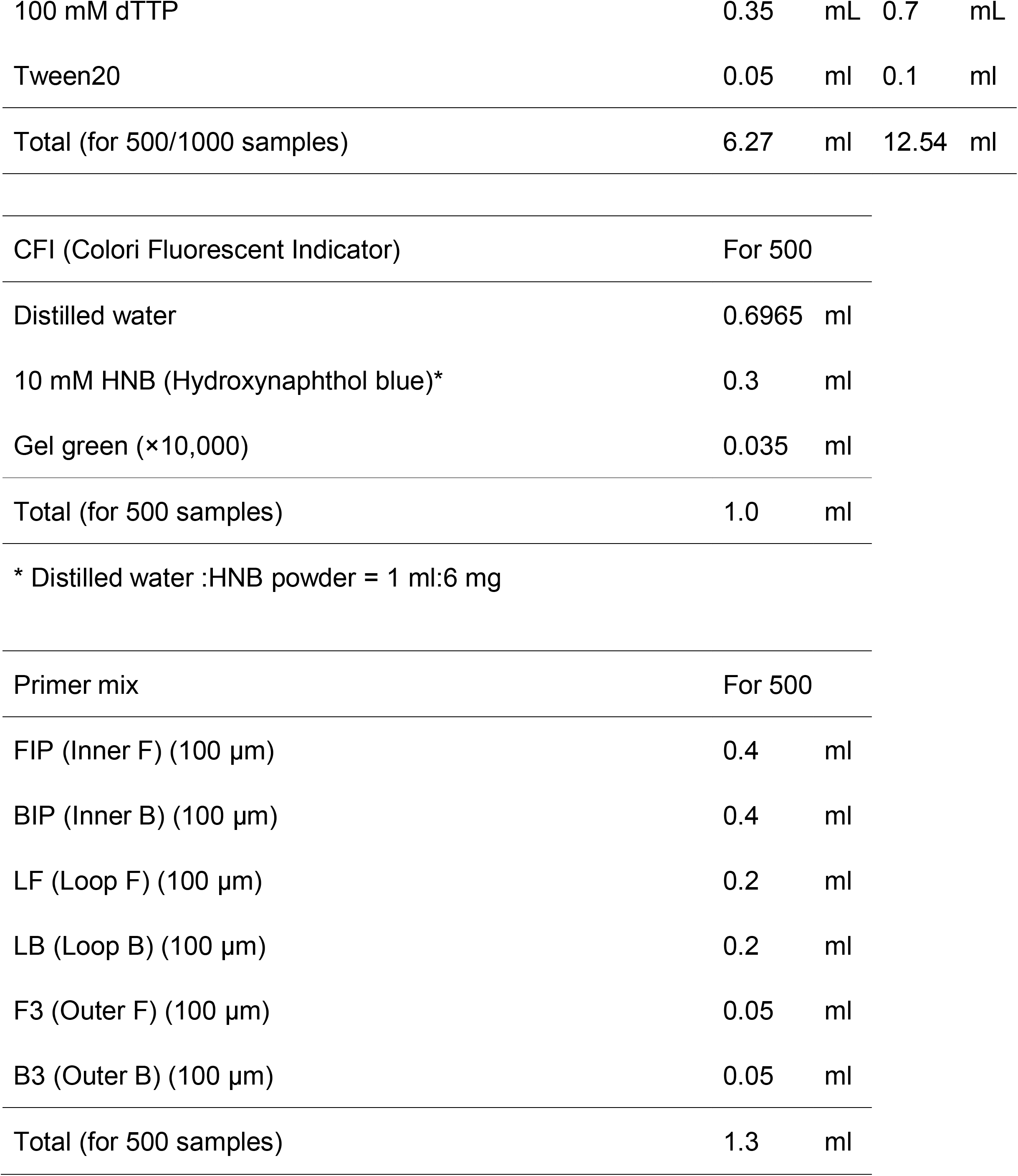
Composition of the RT-LAMP reaction mixture including the homemade master mix.

| RT-LAMP reaction mix | Triplex |  |
| --- | --- | --- |
| 4x LAMP Master Mix | 12.5 | μl |
| Distilled water | 11.5 | μl |
| 1x Primer mix for 1ab | 2.6 | μl |
| 1x Primer mix for S | 2.6 | μl |
| 1x Primer mix for 7a | 2.6 | μl |
| CFI | 2 | μl |
| *Bst* polymerase DNA Large Fragment (8 units/μl) | 6 | μl |
| AMV Reverse Transcriptase for Genie III (20 units/μl) | 0.2 | μl |
| *RNA template* | 10 | μl |
| Total | 50 | μl |

| Homemade 4x LAMP master mix | For 500 |  | For 1000 |  |
| --- | --- | --- | --- | --- |
| 5 M Betaine | 4 | ml | 8 | ml |
| 1 M Tris-HCl (pH8.8) | 0.5 | ml | 1 | ml |
| 3 M Potassium chloride (KCl) | 0.08 | mL | 0.16 | mL |
| 5.6 M (NH<sub>4</sub>)<sub>2</sub>SO<sub>4</sub> | 0.04 | mL | 0.08 | mL |
| 1 M MgSO<sub>4</sub> | 0.2 | mL | 0.4 | mL |
| 100 mM dATP | 0.35 | mL | 0.7 | mL |
| 100 mM dCTP | 0.35 | mL | 0.7 | mL |
| 100 mM dGTP | 0.35 | mL | 0.7 | mL |
| 100 mM dTTP | 0.35 | mL | 0.7 | mL |
| Tween20 | 0.05 | ml | 0.1 | ml |
| Total (for 500/1000 samples) | 6.27 | ml | 12.54 | ml |
| CFI (Colori Fluorescent Indicator) | For 500 |  |  |  |
| Distilled water | 0.6965 | ml |  |  |
| 10 mM HNB (Hydroxynaphthol blue)* | 0.3 | ml |  |  |
| Gel green (×10,000) | 0.035 | ml |  |  |
| Total (for 500 samples) | 1.0 | ml |  |  |

\* Distilled water :HNB powder = 1 ml:6 mg

|  |  |  |
| --- | --- | --- |
| Primer mix | For 500 |  |
| FIP (Inner F) (100 µm) | 0.4 | ml |
| BIP (Inner B) (100 µm) | 0.4 | ml |
| LF (Loop F) (100 µm) | 0.2 | ml |
| LB (Loop B) (100 µm) | 0.2 | ml |
| F3 (Outer F) (100 µm) | 0.05 | ml |
| B3 (Outer B) (100 µm) | 0.05 | ml |
| Total (for 500 samples) | 1.3 | ml |

**S-Table 3.**
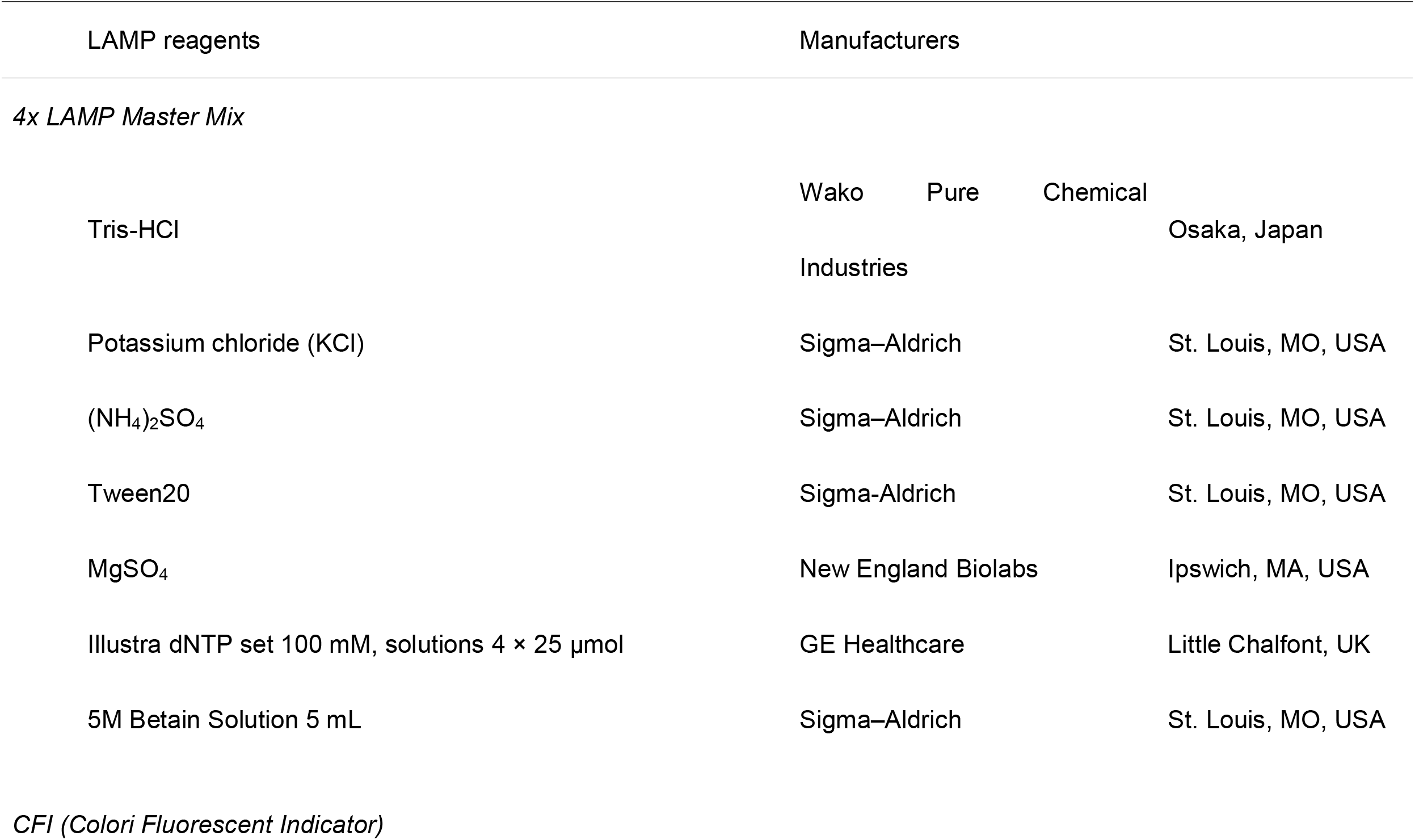
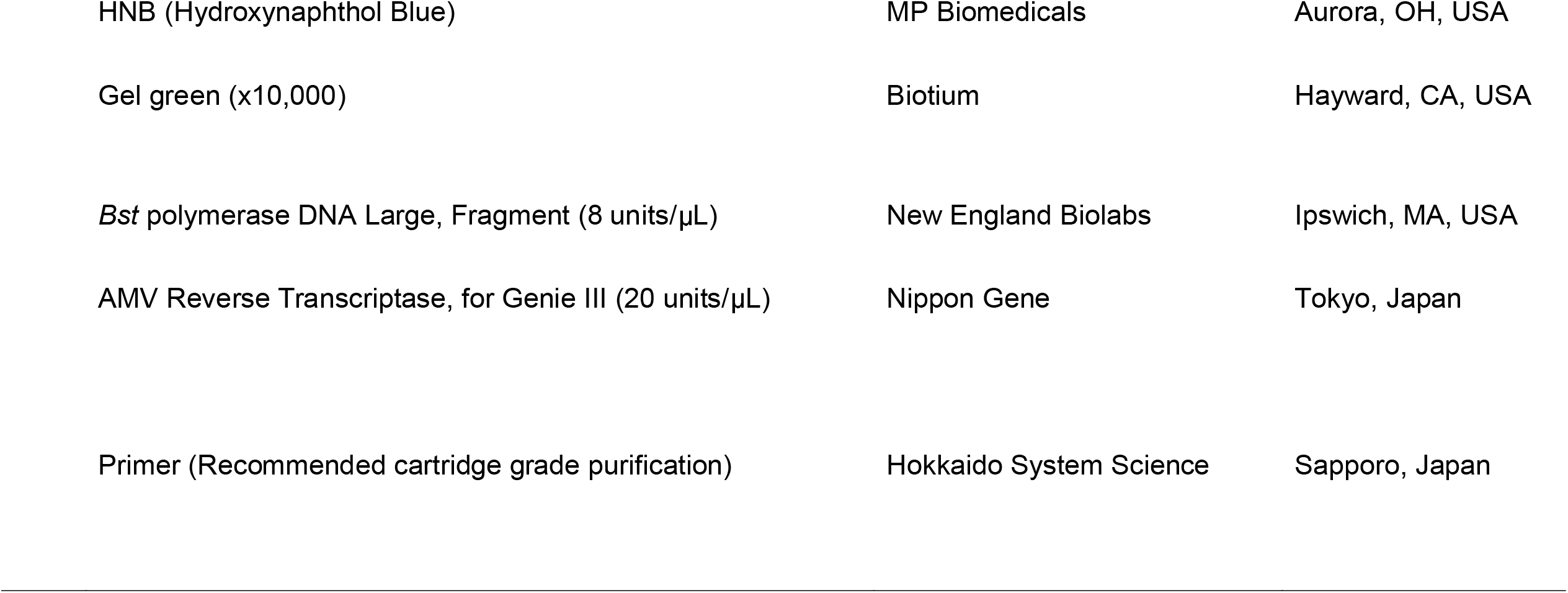
Manufacturers’ list for RT-LAMP reaction mix.

